# Primary progressive aphasia in Italian and English: a cross-linguistic cohort study

**DOI:** 10.1101/2024.03.22.24304752

**Authors:** Salvatore Mazzeo, Chris JD Hardy, Jessica Jiang, Carmen Morinelli, Valentina Moschini, Ella Brooks, Jeremy CS Johnson, Anthipa Chokesuwattanaskul, Anna Volkmer, Jonathan D Rohrer, Assunta Ingannato, Silvia Bagnoli, Sonia Padiglioni, Benedetta Nacmias, Sandro Sorbi, Valentina Bessi, Jason D Warren

## Abstract

**Background and objectives:** Primary progressive aphasia (PPA) signifies a diverse group of neurodegenerative disorders principally affecting language functions. The major syndromic variants of PPA present with distinct profiles of linguistic deficits. However, current concepts and diagnosis of PPA are largely based on English-speaking patients, while few studies have explored how PPA syndromes might vary between languages. Here we undertook a comprehensive neuropsychological comparison of all major PPA syndromes in two languages with contrasting characteristics: Italian and English.

**Methods:** We retrospectively compared the PPA cohorts attending our specialist referral centres on neuropsychological tests sampling a range of linguistic and general cognitive domains. The cohorts comprised 106 native Italian-speakers with PPA (14 nonfluent/agrammatic variant [nfvPPA], 20 semantic variant [svPPA], 41 logopenic variant [lvPPA], 31 mixed PPA [mPPA]) and 166 native English-speakers with PPA (70 nfvPPA, 45 svPPA, 42 lvPPA, 9 mPPA). Neuropsychological scores were normalised to healthy older native speakers (adjusted for age and years of education) and dichotomised (impaired/unimpaired) to identify the proportion of each cohort showing impairment on each test. Cohorts were compared in logistic regression models, covarying for symptom duration and overall cognitive severity.

**Results:** The English PPA cohort was significantly younger (mean 62.7 years) than the Italian cohort (mean 65.9 years; p=0.003), with longer symptom duration (mean 4.6 vs 3.1 years; p=0.048), a higher proportion of nfvPPA cases (42% vs. 13%, p<0.001) and lower proportions of lvPPA (25% vs. 38%, p=0.019) and mPPA (5% vs. 29%, p<0.001). Compared with Italian-speaking patients, English-speaking nfvPPA patients had less frequent expressive agrammatism (p=0.015) and more frequently impaired single-word comprehension (p=0.013) and nonverbal working memory (p=0.041). English svPPA patients had more frequent surface dyslexia (p=0.046) and dysgraphia (p=0.021), while English lvPPA patients had more frequently impaired single-word comprehension (p<0.001), word repetition (p=0.02), nonverbal working memory (p=0.005) and visuospatial perception (p<0.001).

**Discussion:** Language-specific characteristics importantly influence PPA phenotypes: degeneration of language networks may predispose to expressive agrammatism in Italian (reflecting its morphological complexity) and to impaired spoken word processing and regularisation errors in English (reflecting its articulatory, acoustic and orthographic complexity). These findings have implications for diagnosis, management and cross-linguistic collaborative initiatives in PPA.

## 1. INTRODUCTION

Primary progressive aphasia (PPA) designates a diverse group of neurodegenerative disorders led by insidious deterioration of language skills^1^. Current consensus diagnostic criteria for PPA enshrine three canonical syndromic variants based on specific profiles of linguistic impairments^2^: nonfluent/agrammatic variant PPA (nfvPPA), semantic variant PPA (svPPA), and logopenic variant PPA (lvPPA). Diagnostic formulations and our wider understanding of PPA have largely been shaped by studies involving native English speakers^1–3^. This is a serious limitation: languages vary widely in their linguistic, articulatory, acoustic and orthographic characteristics, and neurodegenerative pathologies targeting brain language networks are unlikely a priori to manifest uniformly across languages. This is true even for different languages within the same family. English and Italian, for instance, both belong to the Indo-European language family, but differ widely in linguistic characteristics such as morphological typology, word order, and orthographic depth: whereas English is an analytic language, with few inflections, strict word order and ‘deep’ orthography (i.e., it contains many orthographically irregular words that violate the usual rules of grapheme–phoneme correspondence), Italian is a synthetic language, highly inflected with flexible word order and ‘shallow’ orthography. Further differences extend to the acoustic and articulatory properties of individual speech sounds: for example, English has a greater range of consonantal and in particular vowel phonemes than Italian^4^.

Studies of PPA syndromes in non-English-speaking patients have begun to identify language-specific signatures^5–7^. It is likely a priori that particular linguistic syndromic features of PPA will differ in frequency and characteristics across languages, reflecting underlying variation in essential linguistic properties (for example, the prominence of surface dyslexia will vary between languages, such as Italian and English, with varying orthographic depth)^8^. One study comparing Italian and English-speaking patients with nfvPPA revealed greater motor speech impairment in English-speaking patients and greater grammatical impairment in Italian-speaking patients^9^, after taking into account potentially confounding demographic and clinical factors. These findings are consistent with other work comparing aphasia in Italian and English-speaking patients following stroke^10,11^. However, direct comparisons between PPA syndromes in different languages remain sparse, and we lack detailed cross-linguistic profiles covering the PPA spectrum.

Here we undertook a detailed, retrospective comparison of neurolinguistic and general cognitive profiles in two large, well-characterised patient cohorts comprising Italian and English native speakers with all major syndromes of PPA. We used standardised, dichotomised (impaired/non-impaired) test scores as common metrics with which to compare cognitive domains between cohorts. We hypothesised that the impairment profiles of the two cohorts would differ based on the interaction of native language features with core neurolinguistic domains implicated in canonical PPA syndromes. More specifically, we predicted that the highly inflected nature of Italian grammar would lead to more frequent grammatical errors in Italian speakers with nfvPPA, whereas the greater phonetic demands of English would be reflected in more frequent errors on tests requiring phonetic transformation (word and non-word repetition) in English speakers with nfvPPA and lvPPA, and the greater orthographic depth of English would promote surface dyslexia and dysgraphia in svPPA.

## 2. MATERIALS AND METHODS

### 2.1. Characteristics of the patient cohorts

Italian-speaking patients were recruited and assessed at the Research and Innovation Centre for Dementia, Azienda Ospedaliero-Universitaria, Florence, and English-speaking patients at the Dementia Research Centre, University College London. All were diagnosed according to current international consensus criteria^2^, supported by a comprehensive clinical assessment and neuropsychological evaluation of language and other cognitive domains, and brain MRI and/or ^18^F-deoxyglucose-PET showing a compatible profile of regional atrophy and/or hypometabolism. Patients were classified as having mixed PPA (mPPA) if they fulfilled the criteria for PPA but did not fulfil the criteria for a single canonical PPA variant.

The native English-speaking cohort (henceforth, the ‘English cohort’) comprised 166 consecutive patients with PPA (70 [42%] nfvPPA, 45 [27%] svPPA, 42 [25%] lvPPA, nine [5%] mPPA), recruited between 2005 and 2022. The native Italian-speaking cohort (henceforth, the ‘Italian cohort’) comprised 106 consecutive patients with PPA (14 [13%] nfvPPA, 20 [19%] svPPA, 41 [39%] lvPPA, 31 [29%] mPPA), recruited between 2007 and 2022. Across cohorts, only one patient was bilingual (syndromic diagnosis: mPPA; languages: Italian and French). 125 English-speaking and 94 Italian-speaking patients underwent research genetic screening for known pathogenic mutations in the *GRN*, *MAPT*, *C9orf72*, *APP*, *PSEN1* and *PSEN2* genes.

All analyses comparing the two cohorts were undertaken retrospectively. We excluded patients with a history of significant intercurrent neurological and/or systemic disease, prominent initial episodic memory, visuoperceptual or behavioural impairment, or severe language impairment precluding collection of speech samples of length and quality sufficient for meaningful analysis. None of the patients undergoing MRI showed a significant burden of cerebrovascular disease.

### 2.2. Standard protocol approvals, registrations, and patient consent

All participants gave informed consent to take part in the relevant cohort study and ethical approval was granted by the local institutional ethics committee (the UCL-NHNN Joint Research Ethics Committee or the Institutional Review Board of Careggi University Hospital, Florence, Italy, reference 15691oss), in accordance with Declaration of Helsinki guidelines.

### 2.3. Language and general cognitive tests and standardisation procedures

From the research neuropsychological batteries used to characterise PPA syndromes at the Italian and English specialist centres^12,13^, we selected tests to represent a range of linguistic and non-linguistic cognitive domains. To ensure comparability of the neuropsychological protocols between the English and Italian cohorts, tests were selected if: (i) the same test had been used to assess a particular cognitive domain in both cohorts; or (ii) equivalent tests assessing the same cognitive domain were available for both cohorts and performance on those tests could be standardised based on healthy control norms for each language. The tests used in the cohort comparison are listed in Table S1 in Supplementary Material online. We assessed the following linguistic domains: picture naming, phonemic fluency, category fluency, single-word comprehension, sentence comprehension, word repetition, non-word repetition, sentence repetition, sentence production (expressive grammatical construction), reading and spelling. Assessment of literacy skills presents a particular challenge when comparing languages that vary in orthographic depth: here, reading of orthographically irregular words was assessed in both English and Italian, and any regularisation errors due to sounding these words phonetically (surface dyslexia) were recorded, while the spelling tests assessed both orthographically regular and irregular words (further details in Table S1). Non-linguistic cognitive domains comprised verbal working memory, nonverbal working memory, verbal and spatial episodic memory, visuospatial perception, and executive function.

To perform the cohort comparison, raw neuropsychological scores were transformed as follows. For the English cohort, raw scores on each test were first converted to w-scores^14^ based on mean and standard deviation scores for that test in a cohort of 104 age-matched healthy British controls, adjusted for age and years of education using previously described procedures^15,16^ (mean [standard deviation] age 66.9 [6.7] years, education 15.8 [2.7] years). For the Italian cohort, raw scores on each test were first adjusted for age and years of education based on published normative data for the Italian population^17–23^ and then transformed to z-scores based on published means and standard deviations for that test (see Table S1 for details of the norming cohorts). Based on standardised values, all test scores were next dichotomised as ‘impaired’ (z-score (for the Italian cohort) or w-score (for the English cohort) >1.65, equivalent to 5^th^ percentile performance) or ‘not impaired’. The Italian and English cohorts were then compared based on the proportions of cases in each cohort showing a deficit for each cognitive domain.

### 2.4. Statistical analysis

All statistical comparisons between cohorts were performed using IBM SPSS Statistics Software Version 25 (SPSS Inc., Chicago, USA) and the computing environment R 4.2.3 (R Foundation for Statistical Computing, Vienna, 2013). Distribution normality for variables was assessed using the Shapiro-Wilk test. We extracted descriptive statistics to examine the central tendency and variability of the data using means and standard deviations for continuous variables and frequencies or percentages and 95% confidence intervals (95% CI) for categorical variables, respectively. To compare cohorts and syndromic groups, we used t-tests for normally distributed variables and χ^2^ tests for categorical data. In order to estimate the influence of differential disease severity, comparisons of cohort neuropsychological profiles were additionally adjusted for symptom duration and overall level of cognitive impairment (Mini-Mental State Examination [MMSE] score) using logistic regression. We calculated effect sizes using Cohen’s d for normally distributed numeric measures and Cramer’s V for categorical data.

### 2.5. Availability statement

Anonymized data not published within this article will be made available by request from any qualified investigator

## 3. RESULTS

### 3.1 Comparison of general cohort characteristics

General demographic and clinical characteristics of the Italian and English PPA cohorts are summarised in Table 1. Continuous variables were normally distributed.

**Table 1.**
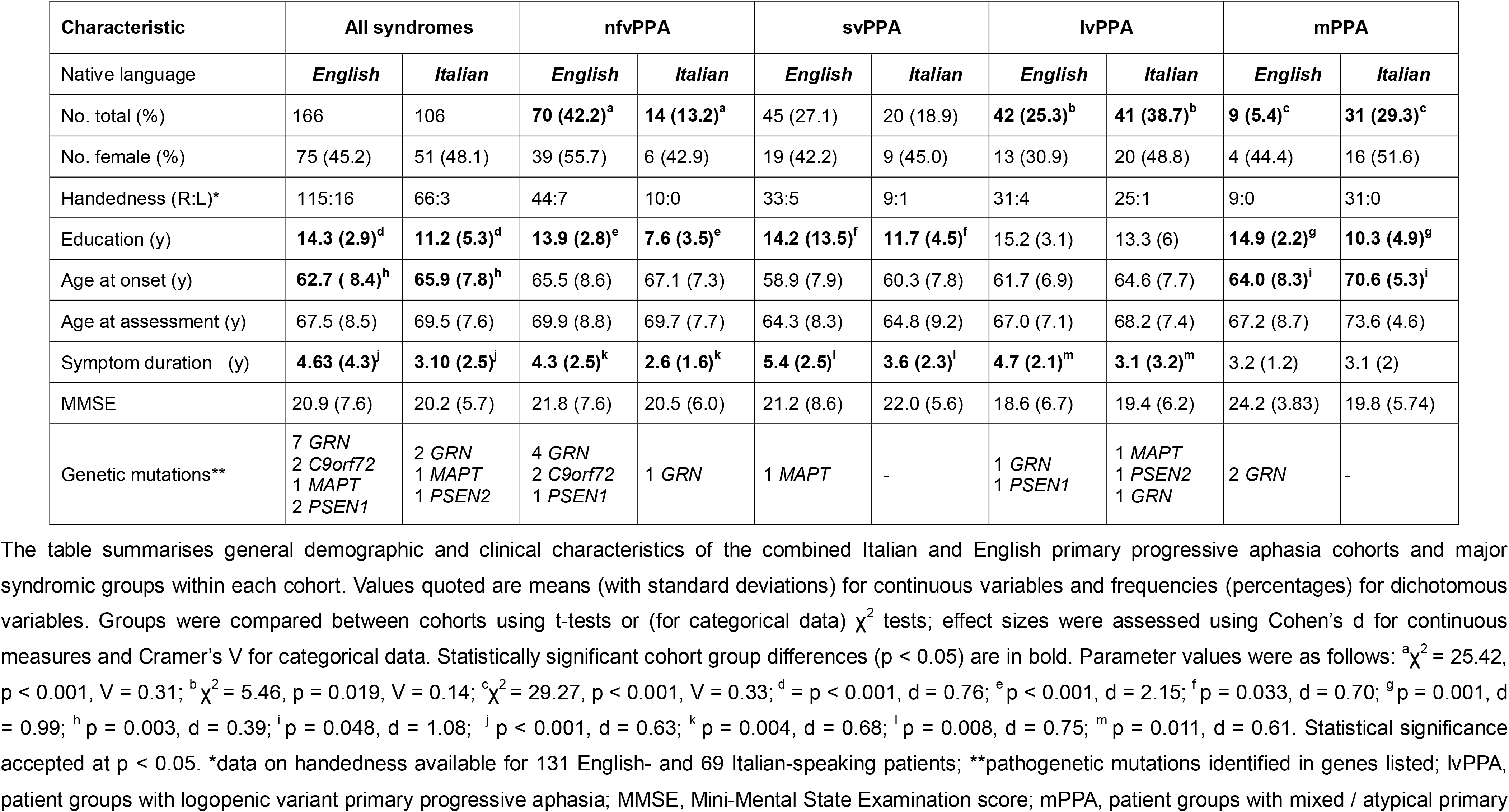
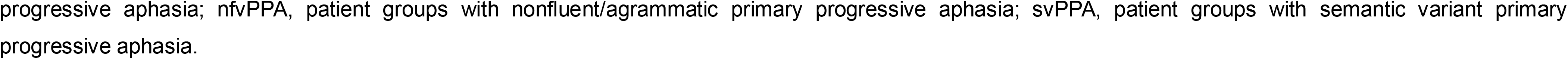
General characteristics of Italian and English primary progressive aphasia cohorts.

The Italian and English cohorts showed no significant differences in sex distribution, handedness, age at assessment or overall cognitive severity (MMSE score). English speakers with PPA had a significantly younger age at onset (p = 0.003, d = 0.39), longer mean symptom duration (p < 0.001, d = 0.63) and more years of education (p < 0.001, d = 0.76) than Italian speakers. Within syndromic groups, similarly directed cohort differences were also observed for symptom duration (nfvPPA, p = 0.004, d = 0.68; svPPA, p = 0.008, d = 0.75; lvPPA, p = 0.01, d = 0.61), education (nfvPPA, p < 0.001, d = 2.15; svPPA, p = 0.03, d = 0.70; mPPA, p = 0.001, d = 0.99) and age at onset (mPPA, p = 0.048, d = 1.08).

The English and Italian cohorts showed distinct profiles of syndromic representation. The English cohort had a higher proportion of nfvPPA (13.2% vs 42.2%, χ^2^ = 25.4, p < 0.001, V = 0.31) and a lower proportion of lvPPA (38.7% vs 25.3%, χ^2^ = 5.46, p = 0.019, V = 0.14) and mPPA (29.3% vs 5.4%, χ^2^ = 29.3, p < 0.001, V = 0.33) than Italian-speaking patients.

Genetic analysis revealed a low frequency of pathogenetic mutations across both cohorts. Mutations in *GRN* were most often represented (nine cases in toto; four English nfvPPA, one English lvPPA, two English mPPA, one Italian nfvPPA, one Italian lvPPA), followed by *MAPT* (two cases; one English svPPA, one Italian lvPPA), *C9orf72* (two cases, both English nfvPPA), *PSEN1* (two cases; one English nfvPPA, one English lvPPA) and *PSEN2* (one Italian lvPPA). Overall, 12/125 (9.6%) of the English cohort and 4/94 (4.2%) of the Italian cohort were found to harbour pathogenetic mutations; these proportions did not differ significantly between the cohorts (p = 0.132).

### 3.2 Comparison of neuropsychological characteristics between cohorts

Neuropsychological data for the two cohorts (English vs Italian) are compared in Table 2, Figure 1 and Figure 2.

**Figure 1.**
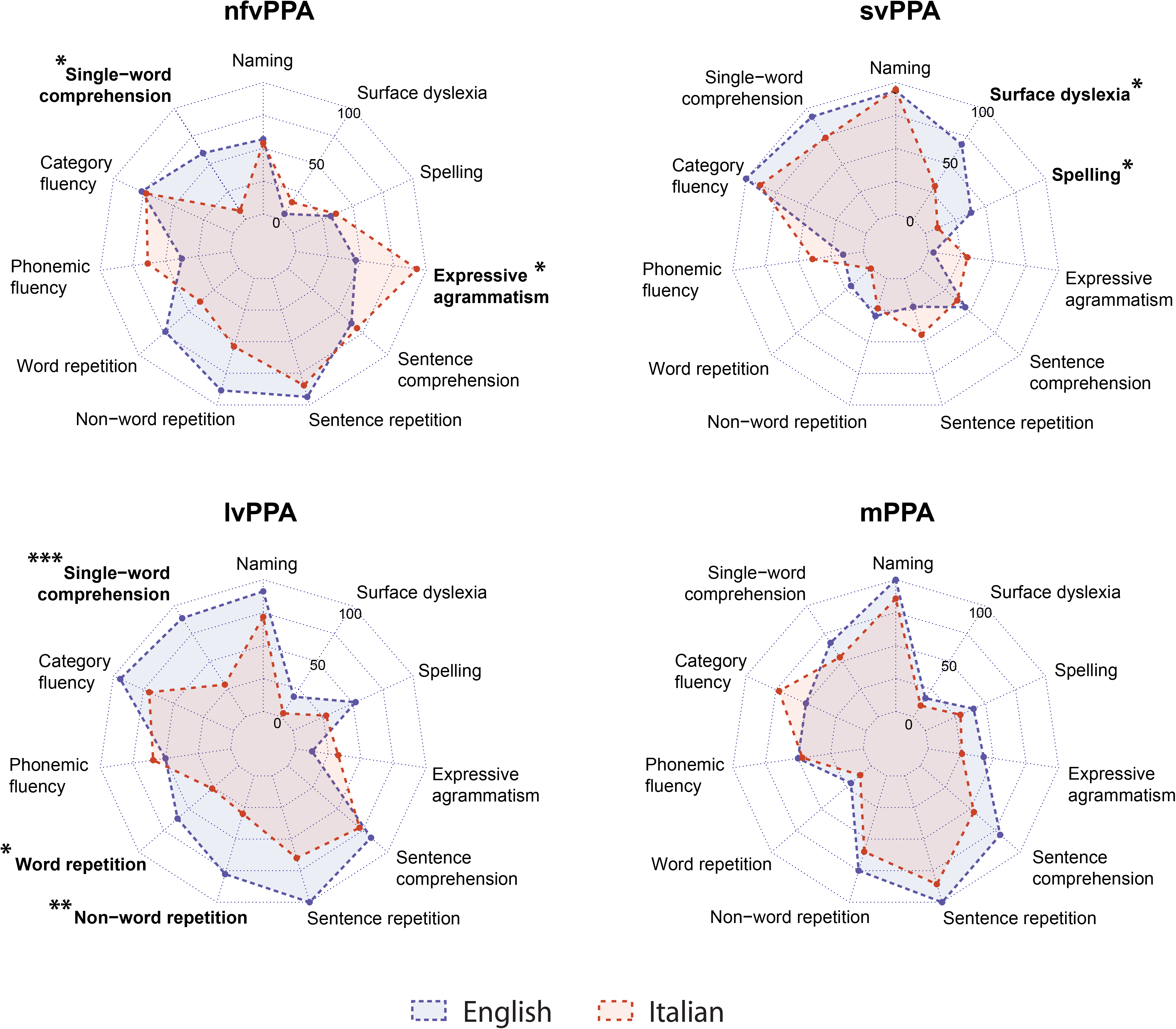
Profiles of neurolinguistic impairment in the Italian and English primary progressive aphasia cohorts. For each major syndromic category, the radar plot represents the proportion of patients showing a deficit in each cognitive domain after z-/w-score transformation (see text and Table 2). Bold characters indicate statistically significant differences between cohorts after adjustment for MMSE and symptom duration, coded as follows: *p < 0.05; **p < 0.01; ***p < 0.001.

**Figure 2.**
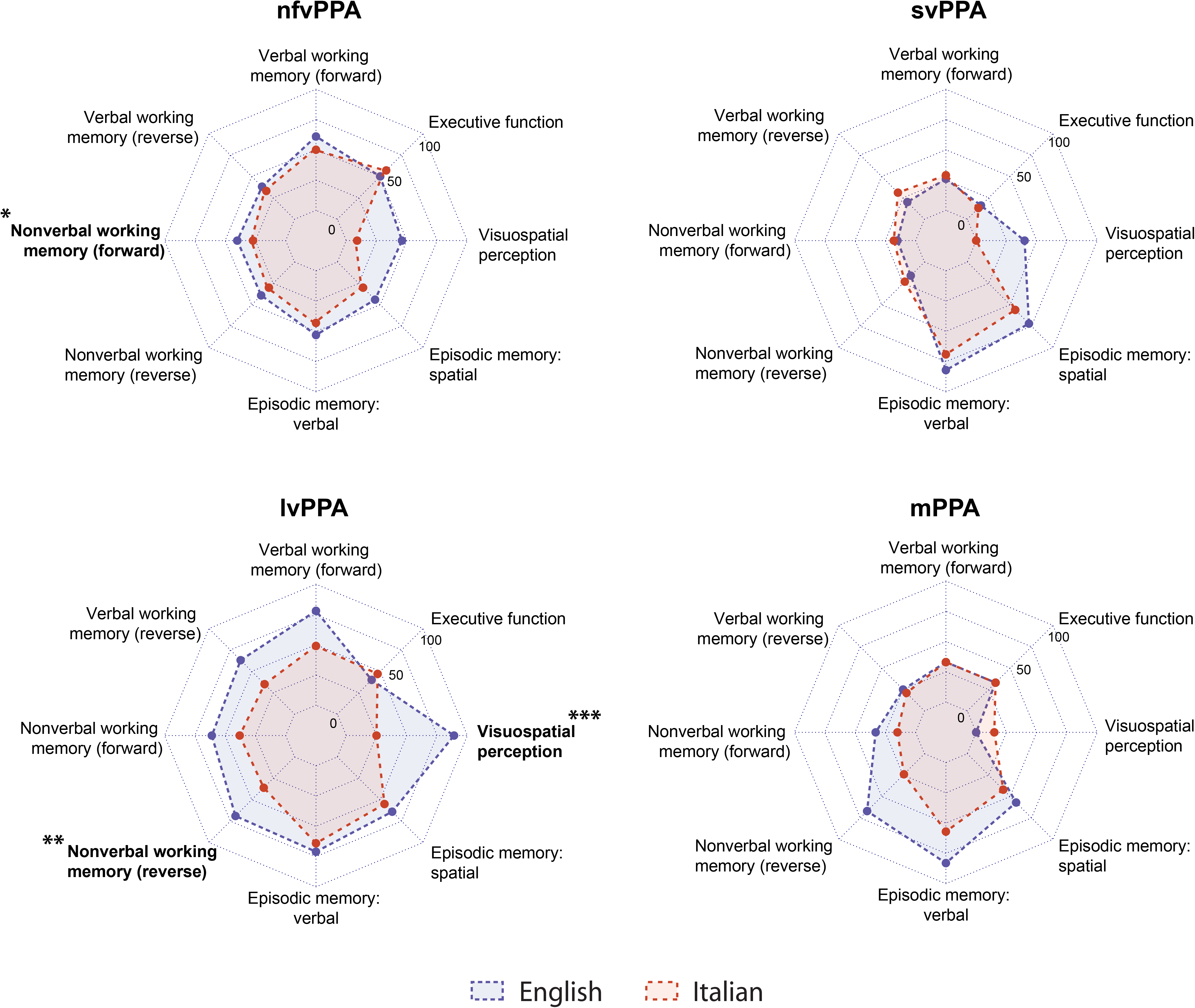
Profiles of non-linguistic general cognitive impairment in the Italian and English primary progressive aphasia cohorts. For each major syndromic category, the radar plot represents the proportion of patients showing a deficit in each cognitive domain after z-/w-score transformation (see text and Table 2). Bold characters indicate statistically significant differences between cohorts after adjustment for MMSE and symptom duration, coded as follows: *p < 0.05; **p < 0.01; ***p < 0.001.

**Table 2.**
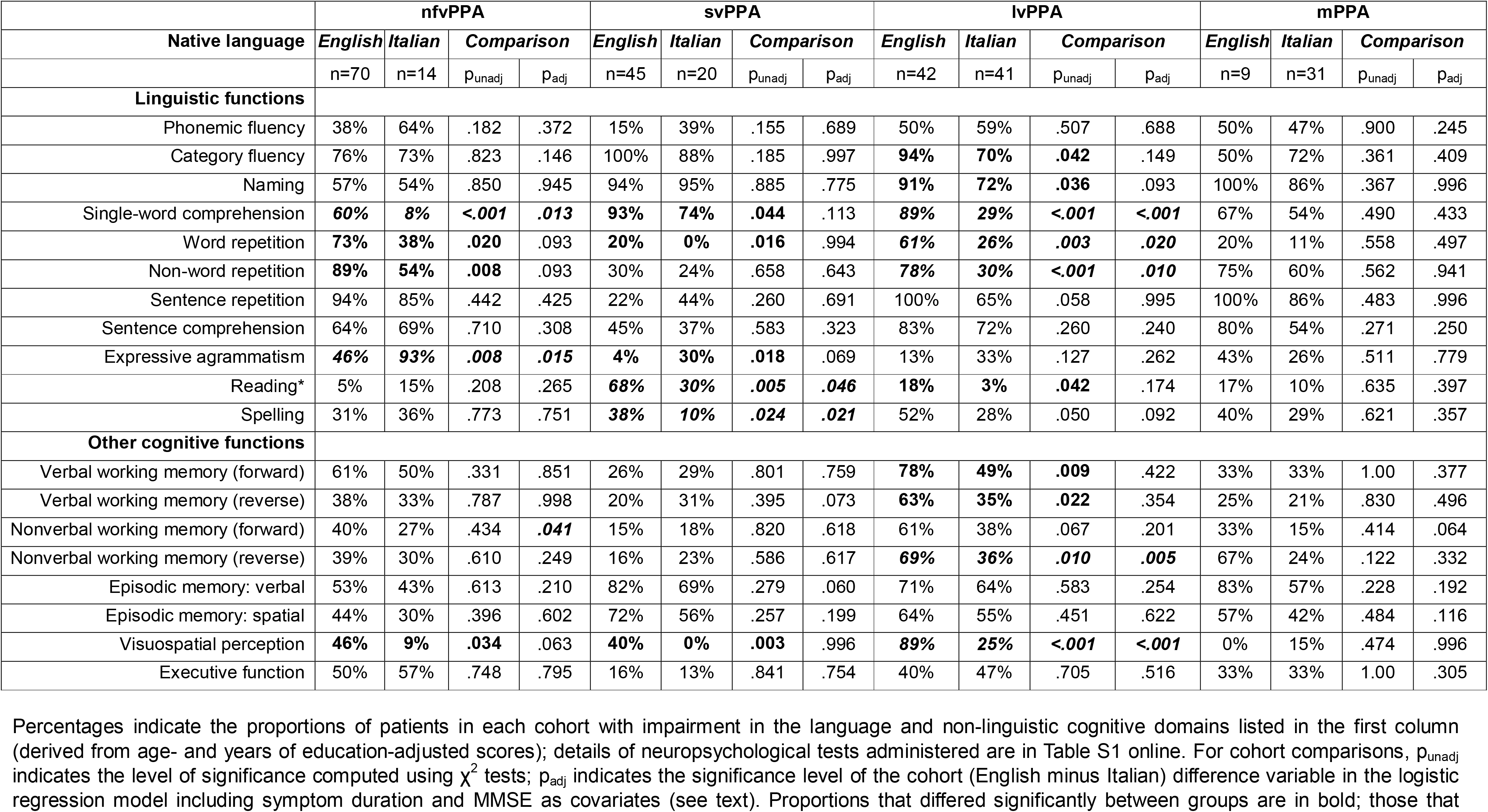
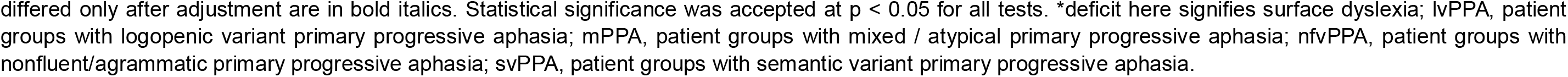
Comparison of the Italian and English primary progressive aphasia cohorts on neuropsychological measures.

|  | nfvPPA |  |  |  | svPPA |  |  |  | lvPPA |  |  |  | mPPA |  |  |  |
| --- | --- | --- | --- | --- | --- | --- | --- | --- | --- | --- | --- | --- | --- | --- | --- | --- |
| Native language | English | Italian | Comparison |  | English | Italian | Comparison |  | English | Italian | Comparison |  | English | Italian | Comparison |  |
|  | n=70 | n=14 | p<sub>unadj</sub> | p<sub>adj</sub> | n=45 | n=20 | p<sub>unadj</sub> | p<sub>adj</sub> | n=42 | n=41 | p<sub>unadj</sub> | p<sub>adj</sub> | n=9 | n=31 | p<sub>unadj</sub> | p<sub>adj</sub> |
| **Linguistic functions** |  |  |  |  |  |  |  |  |  |  |  |  |  |  |  |  |
| Phonemic fluency | 38% | 64% | .182 | .372 | 15% | 39% | .155 | .689 | 50% | 59% | .507 | .688 | 50% | 47% | .900 | .245 |
| Category fluency | 76% | 73% | .823 | .146 | 100% | 88% | .185 | .997 | **94%** | **70%** | **.042** | .149 | 50% | 72% | .361 | .409 |
| Naming | 57% | 54% | .850 | .945 | 94% | 95% | .885 | .775 | **91%** | **72%** | **.036** | .093 | 100% | 86% | .367 | .996 |
| Single-word comprehension | **60%** | **8%** | **<.001** | **.013** | **93%** | **74%** | **.044** | .113 | **89%** | **29%** | **<.001** | **<.001** | 67% | 54% | .490 | .433 |
| Word repetition | **73%** | **38%** | **.020** | .093 | **20%** | **0%** | **.016** | .994 | **61%** | **26%** | **.003** | **.020** | 20% | 11% | .558 | .497 |
| Non-word repetition | **89%** | **54%** | **.008** | .093 | 30% | 24% | .658 | .643 | **78%** | **30%** | **<.001** | **.010** | 75% | 60% | .562 | .941 |
| Sentence repetition | 94% | 85% | .442 | .425 | 22% | 44% | .260 | .691 | 100% | 65% | .058 | .995 | 100% | 86% | .483 | .996 |
| Sentence comprehension | 64% | 69% | .710 | .308 | 45% | 37% | .583 | .323 | 83% | 72% | .260 | .240 | 80% | 54% | .271 | .250 |
| Expressive agrammatism | **46%** | **93%** | **.008** | **.015** | **4%** | **30%** | **.018** | .069 | 13% | 33% | .127 | .262 | 43% | 26% | .511 | .779 |
| Reading* | 5% | 15% | .208 | .265 | **68%** | **30%** | **.005** | **.046** | **18%** | **3%** | **.042** | .174 | 17% | 10% | .635 | .397 |
| Spelling | 31% | 36% | .773 | .751 | **38%** | **10%** | **.024** | **.021** | 52% | 28% | .050 | .092 | 40% | 29% | .621 | .357 |
| **Other cognitive functions** |  |  |  |  |  |  |  |  |  |  |  |  |  |  |  |  |
| Verbal working memory (forward) | 61% | 50% | .331 | .851 | 26% | 29% | .801 | .759 | **78%** | **49%** | **.009** | .422 | 33% | 33% | 1.00 | .377 |
| Verbal working memory (reverse) | 38% | 33% | .787 | .998 | 20% | 31% | .395 | .073 | **63%** | **35%** | **.022** | .354 | 25% | 21% | .830 | .496 |
| Nonverbal working memory (forward) | 40% | 27% | .434 | **.041** | 15% | 18% | .820 | .618 | 61% | 38% | .067 | .201 | 33% | 15% | .414 | .064 |
| Nonverbal working memory (reverse) | 39% | 30% | .610 | .249 | 16% | 23% | .586 | .617 | **69%** | **36%** | **.010** | **.005** | 67% | 24% | .122 | .332 |
| Episodic memory: verbal | 53% | 43% | .613 | .210 | 82% | 69% | .279 | .060 | 71% | 64% | .583 | .254 | 83% | 57% | .228 | .192 |
| Episodic memory: spatial | 44% | 30% | .396 | .602 | 72% | 56% | .257 | .199 | 64% | 55% | .451 | .622 | 57% | 42% | .484 | .116 |
| Visuospatial perception | **46%** | **9%** | **.034** | .063 | **40%** | **0%** | **.003** | .996 | **89%** | **25%** | **<.001** | **<.001** | 0% | 15% | .474 | .996 |
| Executive function | 50% | 57% | .748 | .795 | 16% | 13% | .841 | .754 | 40% | 47% | .705 | .516 | 33% | 33% | 1.00 | .305 |

differed only after adjustment are in bold italics. Statistical significance was accepted at $p < 0.05$ for all tests. \*deficit here signifies surface dyslexia; lvPPA, patient groups with logopenic variant primary progressive aphasia; mPPA, patient groups with mixed / atypical primary progressive aphasia; nfvPPA, patient groups with nonfluent/agrammatic primary progressive aphasia; svPPA, patient groups with semantic variant primary progressive aphasia.

#### 3.2.1 nfvPPA

The English nfvPPA group showed significantly more frequent impairment in single word comprehension (60% vs. 8%, χ^2^ = 11.4, p < 0.001, V = 0.40), non-word repetition (89% vs. 54%, χ^2^ = 6.9, p = 0.008, V = 0.38), word repetition (73% vs 38%, χ^2^ = 5.4, p = 0.020, V = 0.31), and visuospatial perception (46% vs. 9%, χ^2^ = 4.53, p = 0.034, V = 0.36). The Italian nfvPPA group had a significantly higher frequency of expressive agrammatism (93% vs 46%, χ^2^ = 9.20, p = 0.002, V = 0.43). After adjusting for symptom duration and MMSE score, differences in single-word comprehension (p = 0.013), expressive agrammatism (p = 0.015) and nonverbal working memory (p = 0.041) remained significant.

#### 3.2.2 svPPA

The English svPPA group showed significantly more frequent impairment in visuospatial perception (40% vs. 0%, χ^2^ = 9.12, p = 0.003, V = 0.49), reading irregular words (surface dyslexia; 68% vs. 30%, χ^2^ = 7.9, p = 0.005, V = 0.38), word repetition (20% vs. 0%, χ^2^ = 5.9, p = 0.016, V = 0.34), spelling (38% vs. 10%, χ^2^ = 5.1, p = 0.024, V = 0.38) and single word comprehension (93% vs. 74%, χ^2^ = 4.06, p = 0.044, V = 0.26). The Italian svPPA group had a significantly higher frequency of expressive agrammatism (30% vs. 4%, χ^2^ = 5.6, p = 0.018, V = 0.34). After adjusting for symptom duration and MMSE score, differences in spelling (p = 0.021) and surface dyslexia (p = 0.046) remained significant.

#### 3.2.3 lvPPA

The English lvPPA group was significantly more frequently impaired on visuospatial perception (89% vs. 25%, χ^2^ = 18.8, p < 0.001, V = 0.61), single-word comprehension (89% vs. 29%, χ^2^ = 27.3, p < 0.001, V = 0.60), non-word repetition (78% vs. 30%, χ^2^ = 14.7, p < 0.001, V = 0.47), word repetition (61% vs. 26%, χ^2^ = 9.12, p = 0.003, V = 0.35), verbal working memory (digit span forward: 78% vs. 49%, χ^2^ = 6.8, p = 0.009, V = 0.30; digit span reverse: 63% vs. 35%, χ^2^ = 5.2, p = 0.022, V = 0.27), nonverbal working memory (visuospatial span reverse: 69% vs. 36%, χ^2^ = 6.5, p = 0.010, V = 0.33), naming (91% vs. 72%, χ^2^ = 4.4, p = 0.036, V = 0.25), category fluency (94% vs. 70%, χ^2^ = 4.2, p = 0.042, V = 0.27) and reading irregular words (18% vs. 3%, χ^2^ = 4.1, p = 0.042, V = 0.24). The Italian lvPPA group was not significantly more impaired than the English lvPPA group in any neuropsychological domain. After adjusting for symptom duration and MMSE score, differences in visuospatial perception (p < 0.001), single-word comprehension (p < 0.001), nonverbal working memory (p = 0.005), non-word repetition (0.010) and word repetition (p = 0.020) remained significant.

#### 3.2.4 mPPA

The English and Italian mPPA groups did not differ significantly on any of the assessed neuropsychological characteristics at the prescribed threshold.

## 4. DISCUSSION

Here we have shown that the relative frequencies and cognitive profiles of PPA syndromes differ for native speakers of two major word languages, Italian and English, after adjusting for general demographic, disease severity and assessment factors. The frequency of nfvPPA diagnosis was significantly higher in the English cohort, whereas lvPPA and mPPA were higher in the Italian cohort. Genetic mutations were infrequent, occurring in less than 10% of cases in both cohorts. Across syndromes, the English cohort had a higher frequency of specific language deficits than the Italian cohort, though overall cognitive severity was similar in both cohorts. In line with population demographic data^24^ and previous work in PPA^9^, the English cohort on average had a longer education, were younger at symptom onset, and had a longer symptom duration compared to the Italian cohort. Accordingly, we adjusted for each of these factors in analysis, and focus on these adjusted findings below. For nfvPPA, expressive agrammatism was significantly more frequent in the Italian cohort and impaired single-word comprehension in the English cohort; for svPPA, reading (surface dyslexia) and spelling deficits were more frequent in the English cohort; and for lvPPA, single-word comprehension and repetition deficits were more frequent in the English cohort. These profiles are consistent with the grammatical, phonological and orthographic differences between the two languages^4^. Additionally, the cohorts differed in their profiles of non-linguistic cognitive impairment: nonverbal working memory deficits were significantly more frequent in English-speaking patients with lvPPA and nfvPPA, and visuospatial deficits in English-speaking patients with lvPPA.

The relative frequencies of different PPA syndromes are expected to vary somewhat between specialist centres, given that PPA is collectively rare and case ascertainment by any one centre is necessarily limited. However, the relative syndromic proportions in the English PPA cohort here are similar to those reported for previous large British and North American English-speaking PPA cohorts^25,26^. These proportions contrast with the present Italian cohort, particularly in the relative frequencies of nfvPPA, lvPPA and mPPA, while a previous large pan-European study reported nfvPPA and mPPA syndromic frequencies intermediate between the present English and Italian cohorts^27^. Moreover, all cases in both cohorts in this study were diagnosed based on current international consensus criteria^28^. Taken together, this evidence suggests that variations in the syndromic make-up of the Italian and English PPA cohorts here reflect language-based differences rather than merely variable case ascertainment or local diagnostic practices.

Specific differences in PPA syndromic profiles between English and Italian may be relevant here. The diagnosis of nfvPPA rests chiefly on the presence of speech apraxia and expressive agrammatism^2^, and the relative prominence of these features in English and Italian speakers with nfvPPA has previously been shown to differ in line with the present findings^9^. Though we do not have a direct measure of speech sound production errors here, speech apraxia can show language-specific signatures^29^ and may be more likely a priori to manifest in English than in Italian. Whereas English is characterised by complex consonantal clusters that place heavy demands on articulation and a diverse range of vowel sounds, Italian has a simpler syllabic structure and benefits from propulsive prosodic intonation (and relatively fluid syllabic stress) patterns that are close to singing and tend to drive fluency^4,30^. Previous evidence suggests that different error profiles dominate the presentation of speech apraxia in English and Italian^31,32^, with a more diverse range of errors signalling speech apraxia in English speakers. If Italian-speaking patients diagnosed with nfvPPA are less likely to present with speech apraxia, they would be proportionately more likely to present with expressive agrammatism; however, the higher frequency of agrammatism in the present Italian cohort might additionally reflect the contrasting grammatical structures of Italian versus English. Whereas English (as an analytic language) tends to express grammatical relationships through word order and auxiliary verbs, Italian (a highly inflected language) uses a rich set of morphological features that may impose a higher computational load on failing combinatorial mechanisms^33^. In the case of svPPA, the higher frequency of surface dyslexia and dysgraphia in English speakers compared with Italian speakers can be analogously understood as a consequence of the substantially greater prevalence and variety of orthographically irregular words in English^34^. However, the different tests used to assess literacy skills in the two cohorts call for caution in interpreting these deficits.

The greater proportion of single-word comprehension deficits in English-speaking patients with nfvPPA and lvPPA is unlikely to reflect the assessment methods used (since a four-alternative forced choice, spoken word-picture matching task was used in both cohorts^23,35^). A potential neurolinguistic basis for this discrepancy is suggested by the aligned cohort differences in real-word and non-word repetition performance (which attained significance for lvPPA). Relatively greater impairment of auditory phonological decoding in English-speaking patients might lead both to more frequent disruption of auditory working memory and reduced comprehension of spoken words. The greater acoustic complexity of English relative to Italian may be relevant here^4^, and impaired auditory function at or before the stage of phonological decoding has been documented in English speakers with lvPPA and nfvPPA^36–38^. It is noteworthy that impaired single-word comprehension has previously been reported in Dutch-speaking patients with nfvPPA^39^: Similarly to English, Dutch contains frequent, acoustically complex consonant clusters. Further, the discrepancy in spoken word comprehension between cohorts in our study contrasts with their very similar naming performance (see Table 2), arguing for an auditory rather than a primary semantic basis. It is further possible that any phonological input deficit is amplified by concomitantly more marked deficits of phonological production in English-speaking patients with nfvPPA and lvPPA, via impaired lexical predictive processing^40,41^.

The discrepancies in nonverbal function between the Italian and English PPA cohorts may reflect the longer mean illness duration in English-speaking patients; these functions were assessed using the same tests in each cohort, however, the covariates we used to adjust for disease duration and overall cognitive severity (symptom duration, MMSE score) are weighted for verbal deficits. Cohort differences for nonverbal functions were more restricted than for language functions, in line with the argument that specific language characteristics rather than simply diagnostic variability influence the presentation of PPA. This interpretation is bolstered by certain neurolinguistic cohort differences that extended across PPA syndromes: deficits of word comprehension and word repetition were overall more common in the English cohort and expressive agrammatism in the Italian cohort.

The frequency of mPPA was disproportionately higher in the Italian cohort compared to the English cohort. The category of mPPA is nosologically problematic: cases that are unclassifiable under current consensus diagnostic criteria have been described in most large published PPA cohorts, and there remains a lack of consensus on how such cases should be characterised. While mPPA may signify a more advanced disease^42^, it is also recognised that some patients with PPA exhibit language deficits that transcend canonical syndromic boundaries early on^43^. Considering the English and Italian PPA cohorts together, the present study demonstrates that the development of mPPA is not driven by disease duration or severity; indeed, it may manifest on average earlier than canonical PPA syndromes (see Table 1). Further, the neurolinguistic profiles of mPPA cases in the English and Italian cohorts were strikingly similar (Figure 1), arguing for underlying mechanisms that are at least partly common to both language groups. Certain features of the mPPA profile here – in particular the preponderance of naming and sentence repetition deficits – would tend to align it with lvPPA, as previously noted for Italian speakers with mPPA^13^ and in accord with emerging formulations of lvPPA as a multidimensional entity that is loosely demarcated from other PPA syndromes^12,44–46^. The markedly higher frequency of mPPA in the Italian cohort suggests that current diagnostic criteria for PPA^28^, developed for English-speaking patients, may be less well-equipped to encompass the syndromic spectrum of PPA in other languages.

This study has several limitations that should be addressed in future work. Most fundamentally, the English and Italian cohorts did not complete the same neurolinguistic test battery: although we standardised and dichotomised raw scores to mitigate this, the healthy control reference populations also differed, so that standardised scores may not be equivalent between cohorts. Moreover, the sampling of language and other cognitive domains was incomplete (for example, we had no comparison measure of speech apraxia and only assessed a relatively restricted range of nonverbal cognitive skills). Within the cognitive domains sampled, neuropsychological test scores did not align perfectly with the clinical syndromic diagnosis (for example, not all cases receiving a clinical diagnosis of lvPPA in each cohort had impaired scores on naming and sentence repetition; Table 2); this raises the possibility that the clinical application of the diagnostic criteria for PPA may have differed between centres. A more lenient criterion for impaired neuropsychological test scores might have improved their alignment with the syndromic diagnosis but would have tended to make true differences between the cohorts more difficult to detect. A related limitation concerns the use of dichotomised symptom frequency measures to compare cohorts: measures of symptom severity would likely add further information, and this underlines the need for uniform, standardised cognitive tests that can be applied across different language cohorts. Such systematisation will depend on identifying and assessing cognitive equivalents (for example, regularisation errors^47^) in different languages.

The syndromic groups here were relatively small, derived from only two centres and assessed retrospectively; future work should assess larger cohorts prospectively, ideally in the context of a multi-centre collaboration, comparing additional language groups and tracking the longitudinal evolution of deficits (which may also vary between languages). Studies of this kind are already underway^48^ and may be particularly important for rare syndromes such as those associated with genetic PPA: though it is clear that the overall frequency of pathogenetic mutations was low in both cohorts here, this study was not powered to stratify profiles for different mutations. In both sporadic and genetic PPA, there is a need to compare language groups comprehensively – moving beyond neuropsychological tests to assess scales of daily life function, behavioural and neurological features, all of which could potentially vary according to the geographical and ethno-cultural context. Such variation could be amplified by educational differences as well as the population incidence of developmental language disorders^49^ Understanding the neurobiological basis of language-specific PPA signatures will entail neuroimaging, biomarker and ultimately histopathological correlation; this may be particularly pertinent for mPPA^13,26^.

## 5. CONCLUSIONS

Our findings suggest that specific language features may shape the presentation of major PPA syndromes in Italian and English speakers, after adjusting for potentially confounding associations. Specifically, the degeneration of language networks may predispose to expressive agrammatism in Italian (reflecting its morphological complexity) and to impaired spoken word processing and regularisation errors in English (reflecting its acoustic and orthographic complexity). These findings should motivate future large, prospective cross-linguistic studies of PPA with biomarker correlation, to define the nature and extent of syndromic variation across languages. From a neurobiological perspective, such work may illuminate issues (such as the linkage between impaired speech comprehension and production) that are challenging to resolve by studying a single language. The study of fluent bilingual speakers with PPA may be particularly informative^50^. From a clinical perspective, cross-linguistic studies will be essential to develop inclusive, culturally sensitive and appropriate diagnostic guidelines and management interventions for PPA^3,51^ particularly timely in the dawning era of neurodegenerative disease-modifying therapies. From a clinical perspective, cross-linguistic studies will be essential to develop inclusive, culturally sensitive and appropriate diagnostic guidelines and management interventions for PPA^3,51^. As we enter the era of neurodegenerative disease-modifying therapies, cross-linguistic research is of timely importance.

## Acknowledgments

This work was supported by the Alzheimer’s Society, Alzheimer’s Research UK, the UK Dementia Research Institute at UCL, the National Institute for Health Research University College London Hospitals Biomedical Research Centre and RICATEN24 (BN, Ateneo Universita di Firenze, fondi Ateneo 2024). CJDH is supported by a NIHR [Invention for Innovation (NIHR204280)] grant and received a Wellcome Institutional Strategic Support Fund Award (204841/Z/16/Z). JJ was supported by the National Brain Appeal (Frontotemporal Dementia Research Studentship in Memory of David Blechner). CM and VM are supported by a Tuscany Region grant (CUP. D18D20001300002). JCSJ was supported by an Association of British Neurologists Clinical Research Training Fellowship. AV is supported by a NIHR Advanced Fellowship NIHR302240. JDR has been supported by a Miriam Marks Brain Research UK Senior Fellowship, a Medical Research Council Clinician Scientist Fellowship (MR/M008525/1) and the National Institute for Health Research Rare Disease Translational Research Collaboration (BRC149/NS/MH). JDW is supported by the Alzheimer’s Society, Alzheimer’s Research UK, the Royal National Institute for Deaf People (Discovery Grant G105_WARREN) and the National Institute for Health Research University College London Hospitals Biomedical Research Centre. This research was funded in part by UKRI and Wellcome Trust (Grant 204841/Z/16/Z). For the purpose of Open Access, the authors have applied a Creative Commons Attribution (CC BY) public copyright licence to any Author Accepted Manuscript version arising from this submission. The views expressed are those of the authors and not necessarily those of the NIHR or the Department of Health and Social Care.

