## Supplementary materials for "Primary progressive aphasia in Italian and English: a cross-linguistic cohort study"

**Table S1. Neuropsychological tests used to compare English and Italian PPA cohorts**

| **Cognitive domains** | **English cohort** | **Italian cohort** |
| --- | --- | --- |
| **Global cognition** | Mini-Mental State Examination^1^ | Mini-Mental State Examination^2^ |
| **Language functions** |  |  |
| Phonemic fluency | Phonemic fluency task^3^ | Phonemic fluency task^4^ |
| Category fluency | Category fluency task^3^ | Category fluency task^5^ |
| Naming | Boston naming test^6^ | SAND^7^ |
| Single-word comprehension | British picture vocabulary scale^8^ | SAND^7^ |
| Sentence comprehension | PALPA-55^9^ | SAND^7^ |
| Word repetition | Word repetition^10^ | SAND^7^ |
| Non-word repetition | Non-word repetition^10^ | SAND^7^ |
| Sentence repetition | Graded difficulty sentence repetition^11^ | SAND^7^ |
| Expressive agrammatism | Written sentences^12^ | SAND^7^ |
| Reading* | National Adult Reading Test^13^ | SAND^7^ |
| Spelling** | Baxter spelling test^14^ | SAND^7^ |
| **Other cognitive functions** |  |  |
| Verbal working memory | Digit span: forward ^15^ and reverse^3^ | Digit span: forward and reverse^16^ |
| Nonverbal working memory | Corsi blocks: visuospatial span forward and reverse^17^ | Corsi blocks: visuospatial span  forward and reverse^16^ |
| Episodic memory: verbal | Recognition Memory Test words^18^ | Rey Auditory Verbal Learning Test^4^ |
| Episodic memory: spatial | Recognition Memory Test faces^18^ | Rey-Osterrieth Complex Figure Recall^19^ |
| Visuospatial perception | Trail Making Test part A^20^ | Trail Making Test part A^21^ |
| Executive function | Trail Making Test part B^20^ | Trail Making Test part B^21^ |

Only tests for which we administered close equivalents in each cohort are reported. Norms for each test were generated from healthy control cohorts of cognitively normal native speakers. **Control cohort demographic parameters** for tests used were as follows: **English cohort** (**all tests**), n = 104 (59 female), mean (standard deviation) age 66.91 (6.73) years, education 15.81 (2.70) years; **Italian cohort** (varied by test), ***Mini-Mental State Examination***, n = 1019 (769 female), age 75.4 (5.4) years, education 5.2 (2.5) years; ***phonemic fluency***, n = 340 (163 female), age 53.1 (18.0) years, education 10.2 (4.3) years; ***category fluency***, n = 320 (150 female), mean age 47.5 years, mean education 10.1 years (standard deviation values not available); ***digit span*** and ***Corsi blocks***, n = 362 (175 female), age 54.2 (24.1) years, 11.3 (4.6) years; ***Rey Auditory Verbal Learning Test***, n = 340 (163 female), age 53.1 (18.0) years, education 10.2 (4.3) years; ***Rey-Osterrieth Complex Figure Recall***, n = 280 (140 female), age 53.9 (19.8) years, education 11.1 (4.8) years; ***Trail Making Test***, n = 287 (154 female), age 42.2 (15.4) years, education 11.4 (4.7) years; ***other tests***, n = 134 (78 female), age 63.3 (11.2) years, education 11.0 (4.9) years. *In the English cohort, assessed on reading aloud an orthographically irregular word list (the National Adult Reading Test); in the Italian cohort, presence of surface dyslexia recorded on reading aloud irregular words from the SAND. **In the English cohort, assessed on a mixture of orthographically regular and irregular words; in the Italian cohort, assessed on a propositional writing task (patients asked to describe how they brush their teeth); no separate score for surface dysgraphia generated. Abbreviations: F, females; M, males; PALPA-55, Psycholinguistic Assessments of Language Processing in Aphasia subtest 55; SAND, Screening for Aphasia in NeuroDegeneration.
